# Using Generalized Structured Additive Regression Models to determine factors associated with and clusters for COVID-19 hospital deaths in South Africa

**DOI:** 10.1101/2022.09.16.22280020

**Authors:** Innocent Maposa, Richard Welch, Lovelyn Ozougwu, Tracy Arendse, Caroline Mudara, Lucille Blumberg, Waasila Jassat

**Affiliations:** Division of Epidemiology & Biostatistics, School of Public Health, Faculty of Health sciences, University of Witwatersrand; Division of Epidemiology & Biostatistics, Department of Global Health, Faculty of Medicine and Health Sciences, Stellenbosch University; National Institute for Communicable Diseases, Johannesburg, South Africa; Right to Care, Centurion, South Africa

**Keywords:** COVID-19, spatial effects, health systems, hospitalizations, nonlinear effects, clusters, deaths

## Abstract

**Background:** The first case of COVID-19 in South Africa was reported in March 2020 and the country has since recorded over 3.6 million laboratory-confirmed cases and 100 000 deaths as of March 2022. Transmission and infection of SARS-CoV-2 virus and deaths in general due to COVID-19 have been shown to be spatially associated but spatial patterns in in-hospital deaths have not fully been investigated in South Africa. This study uses national COVID-19 hospitalization data to investigate the spatial effects on hospital deaths after adjusting for known mortality risk factors.

**Methods:** COVID-19 hospitalization data and deaths were obtained from the National Institute for Communicable Diseases (NICD). Generalized structured additive logistic regression model was used to assess spatial effects on COVID-19 in-hospital deaths adjusting for demographic and clinical covariates. Continuous covariates were modelled by assuming second-order random walk priors, while spatial autocorrelation was specified with Markov random field prior and fixed effects with vague priors respectively. The inference was fully Bayesian.

**Results:** The risk of COVID-19 in-hospital mortality increased with patient age, with admission to intensive care unit (ICU) (aOR=4.16; 95% Credible Interval: 4.05-4.27), being on oxygen (aOR=1.49; 95% Credible Interval: 1.46-1.51) and on invasive mechanical ventilation (aOR=3.74; 95% Credible Interval: 3.61-3.87). Being admitted in a public hospital (aOR= 3.16; 95% Credible Interval: 3.10-3.21) was also significantly associated with mortality. Risk of in-hospital deaths increased in months following a surge in infections and dropped after months of successive low infections highlighting crest and troughs lagging the epidemic curve. After controlling for these factors, districts such as Vhembe, Capricorn and Mopani in Limpopo province, and Buffalo City, O.R. Tambo, Joe Gqabi and Chris Hani in Eastern Cape province remained with significantly higher odds of COVID-19 hospital deaths suggesting possible health systems challenges in those districts.

**Conclusion:** The results show substantial COVID-19 in-hospital mortality variation across the 52 districts. Our analysis provides information that can be important for strengthening health policies and the public health system for the benefit of the whole South African population. Understanding differences in in-hospital COVID-19 mortality across space could guide interventions to achieve better health outcomes in affected districts.

## BACKGROUND

Over the past two years, the world has grappled with COVID-19, a disease caused by a novel coronavirus, SARS-CoV-2. This was first noted in December 2019 in Wuhan, China, where a cluster of patients with pneumonia of unknown cause was identified (1–3) South Africa recorded its first confirmed COVID-19 case in March 2020 and the government took decisive action to mitigate the spread of the disease through implementing a state of disaster and then adjusted mitigation levels(4) The first wave (peak) of COVID-19 infections occurred in July 2020 in South Africa and four more waves followed (5–8). The most recent waves dominated by Omicron sub-variants have been less severe due to high levels of immunity from vaccination and prior infection in South Africa (5,9,10). As of March 2022, there has been more than 3.6 million confirmed COVID-19 cases and 100 000 confirmed deaths in South Africa, making it the most affected country in Sub-Saharan Africa.

Low- and middle-income countries reported large numbers of COVID-19 related hospitalizations and deaths. New Variants of Concern (VOC) emerged including Alpha, Beta and Delta, that were more transmissible and associated with more severe disease (9). The highly transmissible Omicron BA.1 (B.1.1.529; hereafter BA.1) VOC heralded a new surge of infections in South Africa from November 2021 (10). Two Omicron sub-lineages (BA.4 and BA.5) dominated a surge of new infections from April 2022 (10). The Omicron variant, though highly transmissible, was not as virulent (5)hence had lower hospitalizations and deaths comparably (9,10).

COVID-19 mortality rates were highly disproportionate in South Africa, with the elderly, males, people of colour, those with comorbidities and those admitted at public health facilities and in certain provinces, being at higher risk (6). Several studies have shown that the risk for severe COVID-19 disease were disproportionally born among minority communities (11–13). A South African study highlighted that older age, male sex, minority race groups and lower socioeconomic status (SES) are associated with severe COVID-19 disease and deaths (6). In addition to these factors, patient outcomes generally differ by health sector (public or private hospital) and facility type (district, regional or tertiary hospital) (14).

Jassat et al described the demographic and clinical characteristics of individuals admitted to hospital with laboratory confirmed COVID-19 throughout South Africa in first and second waves using the DATCOV surveillance data and also assessed risk factors for in-hospital mortality(5,6,8). In addition, Jassat et al 2022 described and compared admissions and deaths by age, sex, race and health sector as proxy for SES using same database(6). Waasila et al allude to the role of socio-economic status, race and health care facility type in COVID19 mortality. These can all latently be reflected spatially with similar race groups more likely to be clustered in same neighbourhoods and sharing the same socio-economic status. South Africa being a highly unequal society has a Gini coefficient of 63% (15). Access to efficient health care was reported to be inequitable during the pandemic, and poor communities relied mainly on public health facilities as a consequence of structural inequalities still prevalent in the country (6). The authors also observe a relationship between structural inequality and COVID-19 susceptibility and severity.

The COVID-19 epidemic revealed strong spatial heterogeneity in the spread of infections in countries such as France and Italy (1). A study by Sannigrahi et al 2020 assessed spatial association between socio-demographic variables and COVID-19 cases and deaths and showed distribution of cases and deaths were spatially heterogenous across Europe(16). In England, Sartorius et al 2021 showed that a number of administrative areas (small areas or contiguous small areas) appeared to be at a significantly elevated risk of high COVID-19 transmission and also at increased risk for higher mortality rates(17).

Jassat et al 2022, in South Africa, showed that hospitalized COVID-19 patients in the Eastern Cape and Limpopo provinces were 60% and 50% more at risk of in-hospital deaths respectively compared to those hospitalized in the Western Cape province highlighting spatial heterogeneity in hospital deaths(6). It is evident from these studies that variations in COVID-19 hospital mortality can be found across the socio-economic spectrum with vulnerable communities being at highest risk. This latter study, although it assessed the regional effect, did not account for spatial autocorrelation and also did not model the spatial effect at a finer resolution which can be helpful in defining interventions and deciding effective public health policy.

In recent years, there has been a growing interest in the application of spatial analysis and modelling techniques as a tool for in-depth understanding of public health problems including identifying hotspots, spatial distribution, patterns and effects (1,16–20). SARS-CoV-2 transmission and infections, and COVID-19 deaths, have been shown to be spatially distributed in Europe(16), England(17), and France(1). In Afghanistan, COVID-19 cases were shown to be spatially distributed (21). However, in South Africa, though several studies have been done to determine factors associated with hospital COVID-19 mortality (6), none have considered simultaneously modelling spatial effects, fixed effects and nonlinear effects in investigating factors associated with COVID-19 hospital deaths. In addition, to the best of our knowledge, none of the studies have used flexible structured additive logistic regression models within the Bayesian framework to estimate district spatial effects adjusting for other known COVID-19 mortality risk factors. This study seeks to determine 1) the district level clusters and spatial effects or variability of COVID-19 hospital deaths and 2) identify other factors associated with hospital deaths adjusting for spatial correlation.

## METHODS

### Source of data and sample

The data used in this study was obtained from the NICD. The data was collected using DATCOV an active national COVID-19 hospital surveillance system. The database contains 486 344 hospitalizations with corresponding minimal key individual level data points including age, month and year of hospitalization, patient gender and clinical markers like being on ventilation, on oxygen and admission in ICU. The facility, subdistrict, district and province of the hospital where the patient was admitted are also recorded. We assessed district level spatial effects together with other important covariates that were chosen on the basis of biological plausibility and available evidence. These spatial effects need to be modelled and estimated simultaneously with linear and possibly nonlinear effects. We used district level effects to allow for spatial correlation and any other unknown regional heterogeneity of COVID-19 hospital deaths.

### Bayesian Structured Additive Logistic Regression Model

Let *y*_*ij*_ be the hospital death status for a hospitalized patient *i* in desctrict. *y*_*ij*_ = 1 if the patient *i* in district *j* died in hospital and *y*_*ij*_ = 0 otherwise. A vector *X*_*ij*_ *=* (*x*_*ij*1_, *x*_*ij*2_,*…, x*_*ijp*_)′ contains *p* continuous covariate random variables and *Z*_*ij*_ *=* (*z*_*ij*1_, *z*_*ij*2_,*…, z*_*ijr*_)′ contains some r categorical variables. In our study, *p*= 2 and *r*= 4.

This study assumes that the dependent variable, *y*_*ij*_ is a Bernoulli distributed random variable with *y*_*ij*_ |*p*_*ij*_ *∼ Bernoulli* (*p*_*ij*_) with an unknown *E* (*y*_*ij*_) *= p*_*ij*_, being related to the covariates through the link function

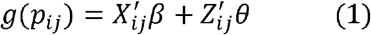

The link function in this equation is known as the logit link, *β* is the *p* dimensional vector of coefficients for the continuous random variables, and *θ* is an *r* dimensional vector of coefficients for categorical random variables. In order to assess for both non-linear effects of continuous random variables and spatial autocorrelation in our data we employed a semi-parametric model which utilizes a penalized regression approach(22,23). The penalized regression approach is a non-parametric method of ordinary least squares (OLS) which relaxes the highly restrictive linear predictor for a versatile semi-parametric predictor. The flexible semi-parametric predictor is defined by:

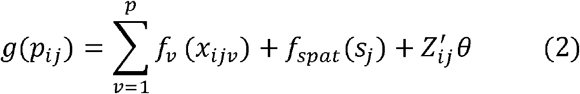

where *f*_*v*_ (·) represents the non-linear twice differentiable smooth function for the continuous covariates and *f*_*spat*_ (*s*_*j*_) is the variable that denotes the spatial effects for each district. In our study, as in Ngesa et al(24), we consider a convolution approach to the spatial effects. The assumption is that the spatial effects can be decomposed into two pure components, that is, spatially structured and spatially unstructured effects given as 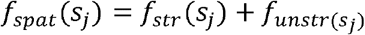. The final model for our study then becomes:

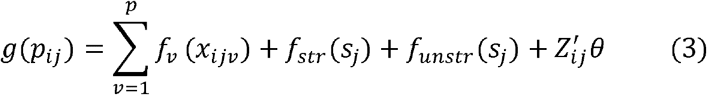

We fit a generalized structured additive logistic regression model for COVID-19 hospital deaths using Markov chain Monte Carlo (MCMC) simulations.

### Data analysis

Statistical analysis was performed in R/RStudio v 4.1.0. Spatial distribution and patterns including Global Moran I and local Moran I for assessing autocorrelation and local clustering were assessed for district level aggregated rates. A conditional autoregressive (CAR) generalized structured additive logistic regression model with binomial link was fit using BayesX R package accounting for spatial effects (25). In addition, non-linear effects of hospital COVID-19 deaths for some continuous covariates were also assessed. The structured additive logistic regression fits a multivariable model with patient sex, facility type, having been on oxygen, ventilator or in ICU as fixed effects, patient age and month (proxy for temporal COVID-19 evolution) as non-linear effects and district as spatial effects. The analysis included data from all waves up to March 2022. All tests were two-sided and a p-value of less or equal to 0.05 was considered to indicate statistical significance. The 95% credible intervals were reported with adjusted Odds Ratios for the full Bayesian inference.

Ethical approval was obtained from the Human Research Ethics Committee (Medical) of University of the Witwatersrand for the DATCOV surveillance programme (M2010108).

## RESULTS

Our analysis included data from all provinces in South Africa over the period March 2020 to March 2022. The database contained 484 699 COVID-19 patients who were hospitalized in either private or public health care facilities. The patient data including hospitalization facility was reported at the individual and subdistrict level. The median age for patients who died was 63 years (IQR: 53-73) while for those who survived or were discharged alive was 48 years (IQR: 34-62). Over half of the patients, 60.9% (n=62928), who died in hospital were admitted and received care from public health care facilities (Table 1).

**Table 1:** COVID19 hospitalized patient characteristics and outcomes, South Africa, March 2020 – March 2022

| Characteristics | Alive n (%) | Died n (%) | p-value |
| --- | --- | --- | --- |
| Gender |  |  |  |
| Female | 215529 (56.5%) | 53131 (51.4%) | <0.001 |
| Male | 165761 (43.4%) | 50278 (48.6%) |  |
| Age (years) | 48 (IQR: 34-62) | 63 (IQR: 53-73) | <0.001 |
| Health sector |  |  |  |
| Private | 191008 (50.1%) | 40481 (39.2%) | <0.001 |
| Public | 190282 (49.9%) | 62928 (60.9%) |  |
| Ever on Oxygen |  |  |  |
| No | 244859 (64.2%) | 47411 (45.9%) | <0.001 |
| Yes | 136431 (35.8%) | 555998 (54.1%) |  |
| Ever in ICU |  |  |  |
| No | 349574 (91.7%) | 72695 (70.3%) | <0.001 |
| Yes | 31716 (8.3%) | 30714 (29.7%) |  |
| Ever ventilated |  |  |  |
| No | 371387 (97.4%) | 84741 (82.0%) | <0.001 |
| Yes | 9903 (2.6%) | 18668 (18.0%) |  |
| Waves |  |  |  |
| Wave 1 | 89398 (23.5%) | 22455 (21.7%) | <0.001 |
| Wave 2 | 90115 (23.6%) | 32537 (31.5%) |  |
| Wave 3 | 118522 (31.1%) | 39596 (38.3%) |  |
| Wave 4 | 83255 (21.8%) | 8821 (8.5%) |  |
| Province |  |  |  |
| Eastern Cape | 32439 (8.5%) | 13224 (12.8%) | <0.001 |
| Free State | 23907 (6.3%) | 6088 (5.9%) |  |
| Gauteng | 118658 (31.1%) | 30288 (29.3%) |  |
| Kwazulu-Natal | 65658 (17.2%) | 17417 (16.8%) |  |
| Limpopo | 14239 (3.7%) | 5293 (5.1%) |  |
| Mpumalanga | 16283 (4.3%) | 4895 (4.7%) |  |
| North West | 26307 (6.9%) | 4962 (4.8%) |  |
| Northern Cape | 8465 (2.2%) | 2444 (2.4%) |  |
| Western Cape | 75338 (19.8%) | 18798 (18.2%) |  |

District level spatial distribution of hospital mortality rates are described in Figure 1 below. Figure 1A shows the distribution of observed crude COVID-19 hospital deaths rates. Most districts in the Eastern Cape including Amathole, Buffalo City, O.R. Tambo, Joe Gqabi, Chris Hani, Cacadu, Nelson Mandela, Alfred Nzo and those in the Limpopo province such as Vhembe, Capricorn, Mopani and Greater Sekhukhune showed elevated observed rates of COVID-19 hospital deaths. Districts such as Xhariep, Mangaung, John Taolo Gaetsewe, Bojanala, Dr Kenneth Kaunda and Sisonke had low observed rates of COVID-19 hospital deaths. Figure 1B shows the significant hot and cold spots (clusters) for hospital COVID-19 deaths using local indicators of spatial autocorrelation (LISA). Most districts in the Eastern Cape show above expected high rates of hospital deaths in neighbouring areas and similar pattern also observed in some districts (eg Mopane) in Limpopo province.

**Figure 1:**
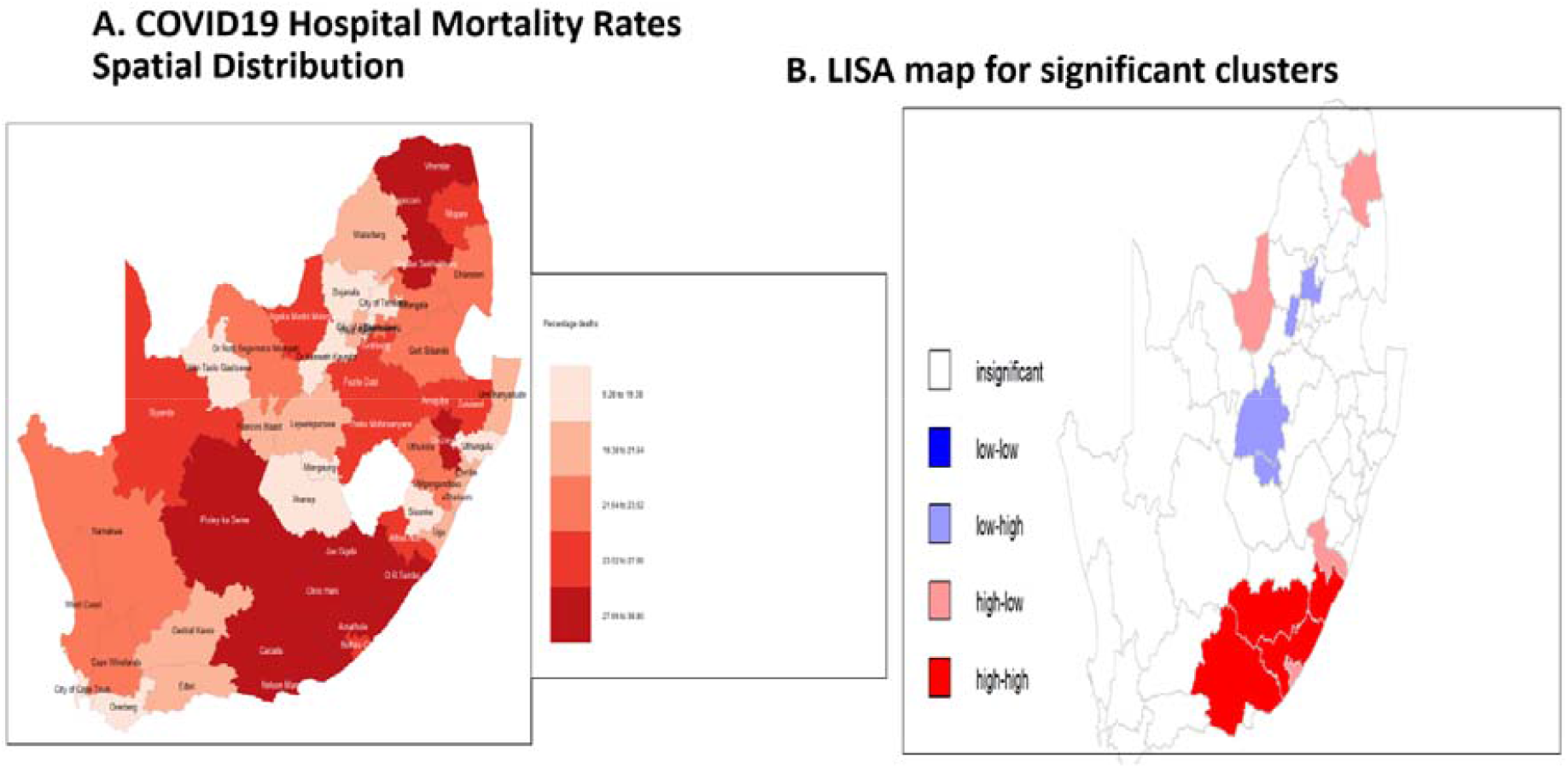
Hospital COVID-19 deaths rates distribution, South Africa, March 2020 – March 2022

Table 2 shows patient and clinical care factors associated with COVID-19 deaths adjusting for period of hospitalization (month) and patient age as nonlinear effects and district level spatial autocorrelation. Adjusting for spatial correlation and nonlinear effects, male gender (adjusted odds ratio [aOR]=1.22; 95% Credible Interval [CI]: 1.20-1.24), admission in a public hospital (aOR=3.17; 95% CI: 3.11-3.23), being on oxygen (aOR=1.37; 95% CI: 1.35-1.39), admitted in ICU (aOR=4.29; 95% CI: 4.18-4.40) and invasive mechanical ventilation (aOR=3.51; 95% CI: 3.40-3.64) were all significantly associated with hospital deaths among admitted COVID-19 patients.

**Table 2:** Patient and clinical care factors adjusted effects associated with mortality, South Africa, March 2020 – March 2022

| Characteristics | Adjusted Odds Ratio (aOR) | 95% Credible Intervals (CI) |
| --- | --- | --- |
| Gender |  |  |
| Female | 1 |  |
| Male | 1.22 | 1.20-1.24 |
| Facility Sector |  |  |
| Private | 1 |  |
| Public | 3.17 | 3.11-3.23 |
| Treated with Oxygen |  |  |
| No | 1 |  |
| Yes | 1.37 | 1.35-1.39 |
| Treated in ICU |  |  |
| No | 1 |  |
| Yes | 4.29 | 4.18-4.40 |
| Ventilated |  |  |
| No | 1 |  |
| Yes | 3.51 | 3.40-3.64 |
| Random effects | Estimate | 95% CI |
| Spatially structured effects<br>[$f_{str}(s_j)$] | 0.014 | 0.0007- 0.0536 |
| Spatially unstructured<br>effects [$f_{unstr}(s_j)$] | 0.0672 | 0.0382 – 0.1067 |

### Nonlinear effects of patient age and admission month

Figure 2 shows the nonlinear effect of time in months on the likelihood of hospital COVID-19 deaths in South Africa over the years 2020 to 2022. The figure provides posterior mean of the smooth time function and its corresponding 80% as well as 95% Credible Intervals. It is clear from the figure that the risk of hospital deaths fluctuated significantly over the peak of the COVID-19 waves. The association between time in months and COVID-19 hospital deaths was nonlinear and assuming linear effect would bias the result and lead to incorrect interpretation of time effect.

**Figure 2:**
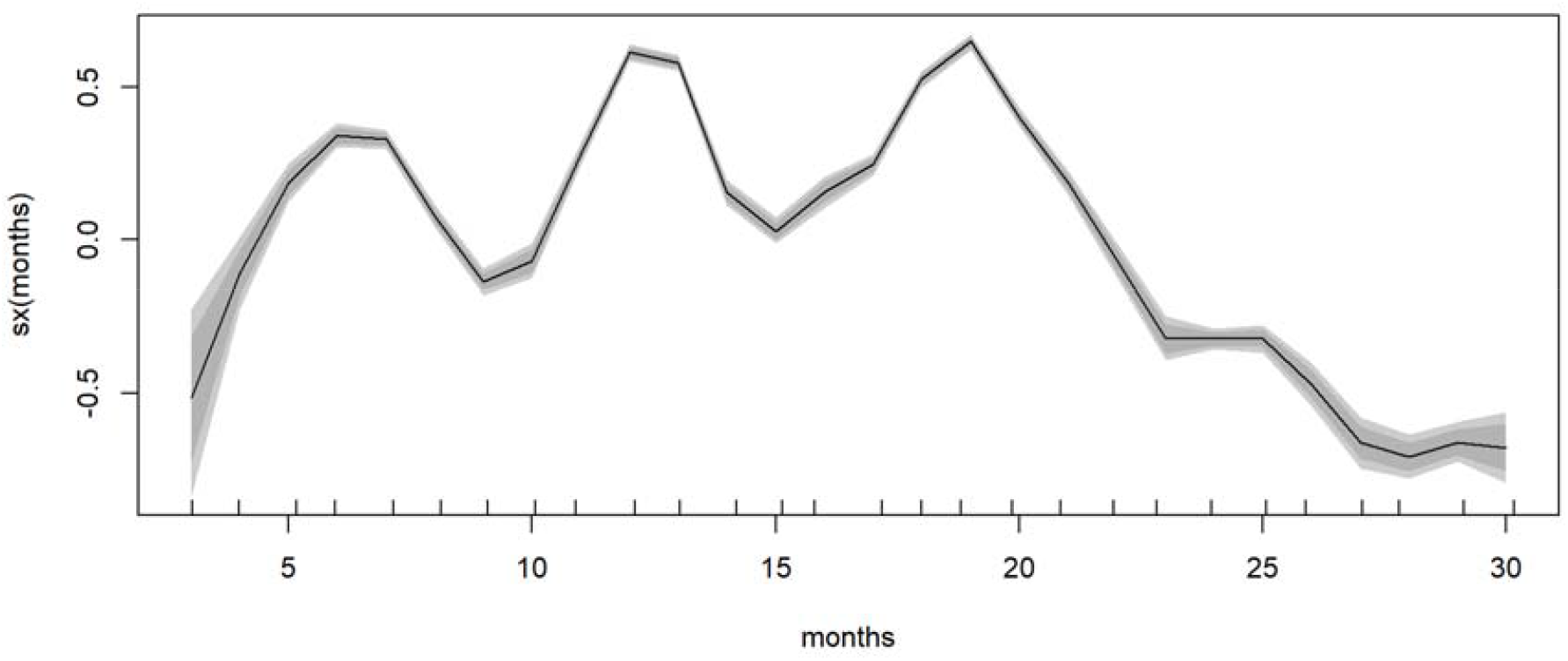
Nonlinear effect of time in months on COVID-19 in-hospital deaths, South Africa, March 2020 – March 2022

Figure 3 shows the nonlinear effect of age on hospital COVID-19 deaths. The likelihood of hospital deaths increased with increasing age in South Africa over the peak periods of the epidemic. The figure provides posterior mean of the smooth age function and its corresponding 80% as well as 95% Credible Intervals. The association between patient age and COVID-19 hospital deaths was nonlinear and assuming linear or peace-wise effect, as with month of hospitalization, would bias the result and lead to incorrect interpretation of age effects.

**Figure 3:**
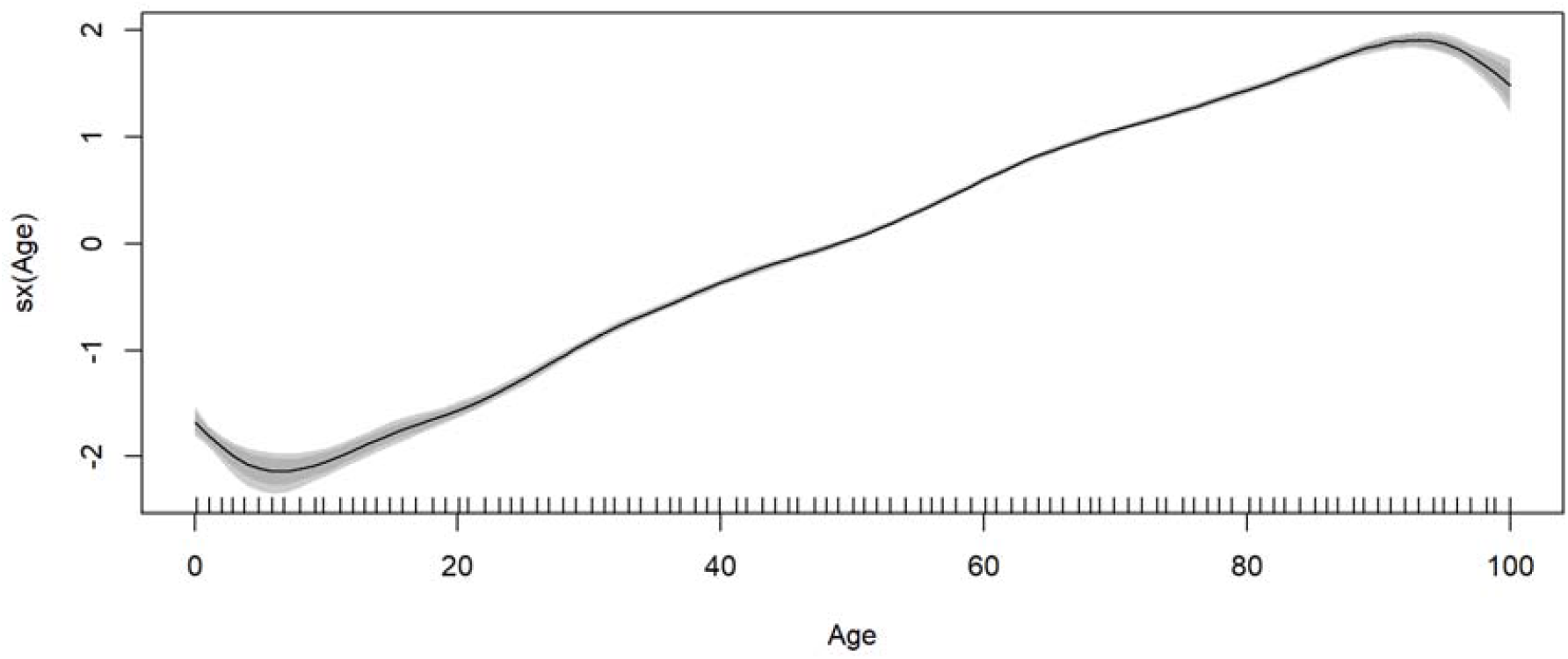
Nonlinear effect of patient age on COVID-19 in-hospital deaths, South Africa, March 2020 – March 2022

We investigated the spatial effects using the structured additive logistic regression model and figure 4 shows the residual spatially associated hospital deaths after adjusting for fixed and nonlinear effects. Districts with blue colour show lower odds of hospital deaths while those with red colour indicate significant higher odds of hospital deaths. From Figure 4, there is clear evidence of spatial variation of hospital COVID-19 deaths after controlling for some known risk factors. The spatial random effect, that is, the heterogeneity between districts in COVID-19 deaths dominates the residual spatial variation explaining about 83% of the variance showing differences in in-hospital COVID-19 deaths by district.

**Figure 4:**
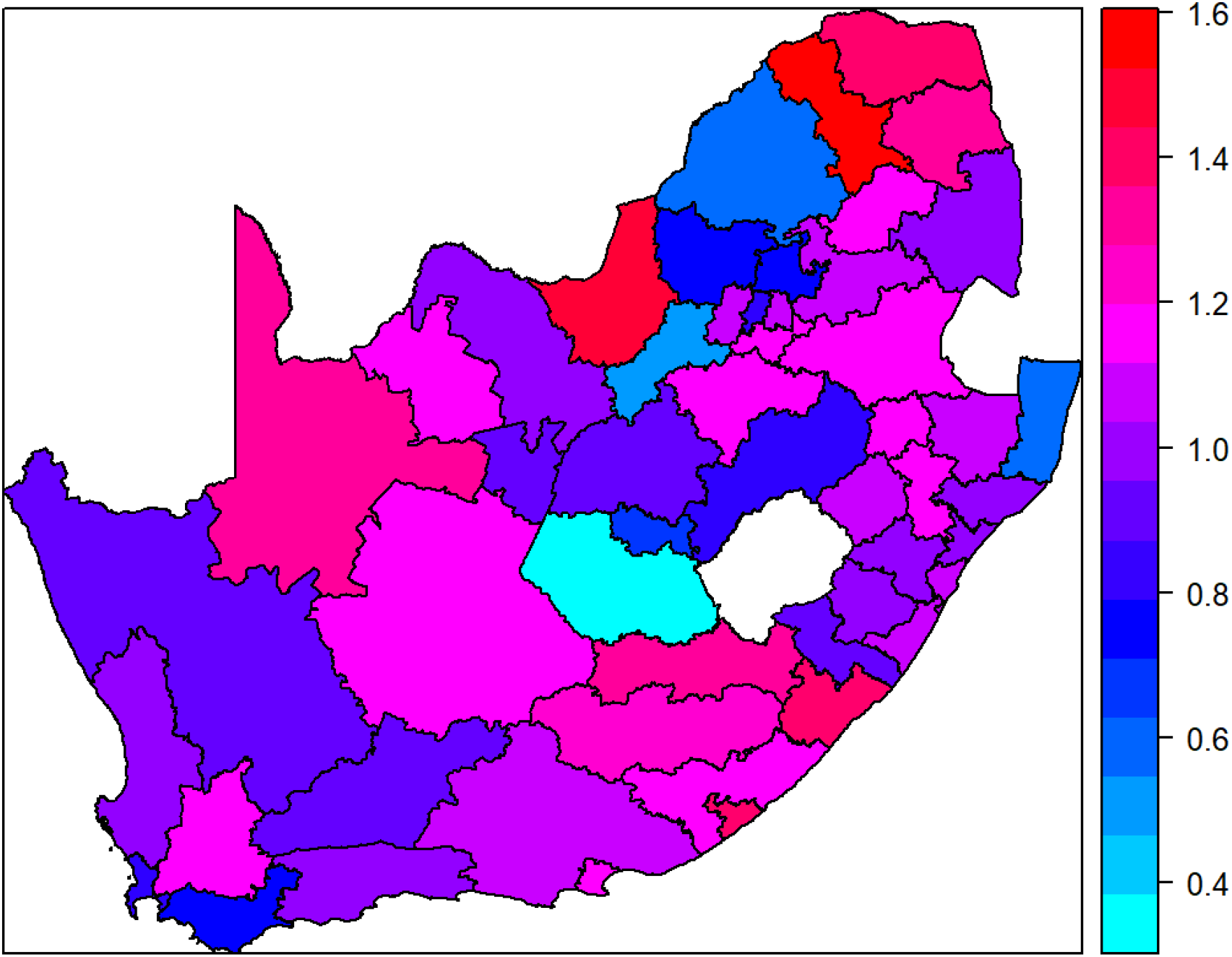
Residual spatial odds of in-hospital deaths, South Africa, March 2020 – March 2022

## DISCUSSION

We examined factors associated with in-hospital COVID-19 mortality in South Africa between March 2020 and March 2022, adjusting for spatial autocorrelation at district level, fixed effects and some non-linear temporal and age effects. Besides confirming that older age, male gender, admission in ICU, being treated with supplemental oxygen and invasive mechanical ventilation, predisposed hospitalized COVID-19 patients to a higher risk of in-hospital mortality, our study highlighted substantial heterogeneity in mortality across districts.

Our study describes the spatial distribution of COVID-19 in-hospital deaths in the whole population in South Africa adjusting for several factors including month of admission as a proxy for epidemic waves. The impact of spatial neighbourhood dependents and heterogeneity have not previously been explored at higher resolution in South Africa regarding COVID-19 in-hospital mortality.

To respond effectively to new epidemics, it is essential to unpack the spatial dynamics of in-hospital deaths at higher spatial resolution such as at subdistrict or district level in order to identify priority areas for intervention to improve health outcomes. Our spatial model adjusted for temporal effects, suggested that age had a nonlinear effect on in-hospital mortality with increasing risk for increasing age as was determined in other previous studies (6,12,26–29).

The study also provides evidence that in-hospital mortality was clustered geographically with several districts in Eastern Cape and Limpopo showing elevated risk. Previous studies showed that hospitalized COVID-19 patients in the Eastern Cape and Limpopo provinces were at higher odds of death (6), however, our study goes further in identifying the districts in those provinces which predisposed patients to higher odds of in-hospital mortality. A study in Brazil similarly explored the spatial distribution of COVID-19 cases and deaths in paediatric population and showed that forty municipalities had higher mortality, and these were observed in regions with poor socio-economic indicators and greatest health disparities (19). Another study in South Africa assessed spatial distribution of deaths at provincial level and other COVID-19 outcomes showed that Eastern Cape had higher risk for deaths (30,31).

Understanding spatial heterogeneity in relation to the socio-environmental determinants and COVID-19 related outcomes is central to targeting interventions for vulnerable populations (1). Our results indicate substantial geographical variations in the distribution of COVID-19 in-hospital mortality across South Africa districts. In South Africa, 21% of the population have access to private health care while the remainder rely on public health care support (6). There are also inequities in health care access in South Africa, with poorer access and availability of health facilities in rural provinces such as Eastern Cape, KwaZulu-Natal and Limpopo. Quality of care has also historically been poor in many public health facilities (14).

A study that assessed inequalities in access to health care in South Africa showed large differentials in private health care admissions between people living in the poorer, more rural provinces like Limpopo, Eastern Cape and Mpumalanga compared to more urban provinces like Gauteng (32). We provide evidence of spatial clustering and spatial heterogeneity in in-hospital mortality in South Africa. Highlighting districts at excess risk of COVID-19 in-hospital mortality can help guide local health care system policies to better protect vulnerable population subgroups. Health care access inequality is not a new aspect in South Africa, however, when it contributes to some groups of the population left at higher risk of in-hospital mortality, this calls for immediate action for implementation of policies that enhance equal access to and improved delivery of health care(6,14,32).

Besides showing spatial structure in the COVID-19 in-hospital mortality in South Africa, our study using the non-linear effects of months as a proxy for temporal evolution of the epidemic highlight higher risk of deaths during the second and third waves. Jassat et al similarly reveal in their study that in-hospital case-fatality ratio during the Omicron wave was 10·7%, compared with 21·5% during the first wave, 28·8% during the second wave, and 26·4% during the third wave (7,8,33). The Beta and Delta VOCs drove the second and third COVID-19 waves in South Africa. Our study reveals an ebbing off in the in-hospital deaths during the last months which were driven by the latter variants of Omicron. Omicron marked a change in the SARS-CoV-2 epidemic curve, clinical profile, and deaths in South Africa (33) Davies et al showed similar patterns in the Western Cape province (9).

Major strengths of this study include the application of flexible structured additive logistic regression model within the Bayesian framework which allows for exploration of spatial association with health outcomes at higher spatial resolutions as well as allow for inclusion of non-linear effects and fixed effects. In addition, this study utilizes a COVID-19 hospital admissions high-quality national data repository with their associated individual level outcomes ensuring ample admission sample data as well as outcomes for analysis at district level. This study is not without some limitations. For this analysis, we adjusted for some of the known risk factors for COVID-19 in-hospital deaths, however, there may be other covariates which we may not have included. A further limitation may be that the DATCOV database does not distinguish between patients who were admitted with coincidental positive SARS-CoV-2 test and those with COVID-19. As highlighted in the preceding limitations, there was limited information on other important variables and ancillary predictors that can be used to model in-hospital mortality. Therefore, there is a need to utilize the same modelling framework to account for the impact of these and other factors especially aggregated at subdistrict or district levels.

As in one Brazilian study (19), our findings confirmed the higher burden of COVID-19 in-hospital mortality in some districts in Limpopo and Eastern Cape provinces. This may be linked to poor health care access including limited access to private health care (32) due to unaffordability of medical insurance. To reduce the burden of in-hospital deaths due to potentially new epidemics and COVID-19 in particular, the South African government and private stakeholders need to build strong and resilient health care infrastructure which can be accessed equally by both the poor and rich independent of class or race especially in vulnerable districts. Modelling the effects of underlying factors and in-hospital disease mortality at small spatial scale is essential to plan effective control strategies for disease outcome risks (20). The relationship between COVID-19 in-hospital mortality rates and sociodemographic, clinical and districts spatial dependents can consequently guide the development of specific intervention actions for these places.

## Conclusions

The results from this study reveal substantial COVID-19 in-hospital mortality variation across the districts in South Africa. This highlights the importance of modelling spatial patterns simultaneously with fixed and nonlinear effects of continuous covariates to identify clusters at high risk of health outcome. The flexible approach to modelling data that has spatial patterns helps to account for possible loss of efficiency due to spatial correlation that spatial patterns can induce in data. Our analysis suggests notable COVID-19 hospital deaths clustering in some districts in Limpopo and Eastern Cape provinces and this information can be important in strengthening the public health and health systems policies for the benefit of the whole South African population. Understanding differences in in-hospital COVID-19 mortality across space could guide interventions to achieve better health outcomes.

The findings of this analysis reveal that strengthening health systems, specifically critical intensive care, oxygen support, and availability of health care workers, could lead to substantial reductions in mortality rates in South Africa in the case of a resurgence of COVID-19 epidemic or new epidemics. It is also important for public health authorities to manage medical care effectively and in a timely manner, as well as to direct the intensity and type of interventions needed to overcome the pandemic.

## Data Availability

The dataset analysed for the manuscript is available upon reasonable request. The data dictionary is available at request to the co-author

## ACKNOWLEDGEMENTS

We acknowledge the NICD team responsible for reporting hospitalisation data. Thanks to the National Department of Health and the NICD for support and oversight. Our gratitude to the laboratories, clinicians and data teams in all public and private sector hospitals throughout the country reporting cases and hospitalisation data, who are acknowledged and listed as the DATCOV author group: https://www.nicd.ac.za/diseases-a-z-index/covid-19/surveillance-reports/daily-hospital-surveillance-datcov-report/

